# A Health Economic Evaluation for Implementing an Extended Half-life Monoclonal Antibody for All Infants vs. Standard Care for Respiratory Virus Syncytial Prophylaxis in Canada

**DOI:** 10.1101/2024.03.28.24305020

**Authors:** Thomas Shin, Jason KH Lee, Alexia Kieffer, Michael Greenberg, Jianhong Wu

## Abstract

Respiratory syncytial virus (RSV) is a highly infectious virus, and infants and young children are particularly vulnerable to its progression to severe lower respiratory tract illness (LRTI). Nirsevimab, an extended half-life monoclonal antibody, was recently approved in Canada as a passive immunization intervention for the prevention of RSV LRTI. A static decision tree model was utilized to determine the cost-effectiveness of nirsevimab in Canadian infants compared to current standard of care (palivizumab for infants born preterm, and with specific chronic conditions) and generate an optimal price per dose (PPD) at accepted willingness-to-pay (WTP) thresholds. Various health outcomes (including hospitalization, ICU, and mechanical ventilation) and healthcare costs were calculated over one RSV season, with any necessary follow-up prophylaxis in the second season for three infant categories (palivizumab-eligible, preterm, and term). All health-related parameters and costs were tailored to the Canadian environment. Compared to scenarios where only at-risk segments of the infant population received nirsevimab, the base case (administering nirsevimab to all infants in their first RSV season) was the most cost-effective versus standard care: the PPD was $692 at a $40,000/QALY WTP threshold, using average costing data assumptions across all scenarios. Compared to standard care, the base case scenario could avoid 18,249 RSV-related health outcomes (reduction of 9.96%). Variations in discount rate, distribution of monthly RSV infections, nirsevimab coverage rate for infants born at term, and palivizumab cost had the most significant model impact. Passive immunization of all infants with nirsevimab can significantly reduce RSV-related health and economic burden across Canada.

## Introduction

Respiratory syncytial virus (RSV) is a highly infectious and prevalent viral pathogen with activity that follows an annual seasonal pattern.^1,2^ Afflicting individuals of all ages, RSV is transmitted via respiratory droplets and exhibits a typical presentation of mild upper respiratory tract symptoms such as sneezing, coughing and runny nose that resolve in 1-2 weeks.^3^ However, subpopulations such as infants and young children are particularly vulnerable to more severe illness with RSV,^4^ presenting as acute lower respiratory tract infection (LRTI), most commonly bronchiolitis and pneumonia. In Canada, RSV is the leading cause of hospitalization in children four years and younger,^5^ and 80% of these children were born at term with no underlying health conditions.^6^ While RSV-associated death rates are highest in infants who were born premature, the mortality burden is greatest in full-term infants given their proportionately large population.^7^ For children who recover from RSV, early infection may also have long-lasting health consequences, as indicated by associations with recurrent wheezing and pediatric asthma.^8^ Infants are at the highest risk for severe manifestations of this virus during their first RSV season,^9^ and nearly all children will be infected by two years of age;^10^ this high prevalence, combined with the potential severity and long-term impact of RSV, signifies a tremendous unmet medical need to protect infants from this infection.

RSV-related prophylaxis among infants represents a promising tool for reducing the significant health and economic burden of RSV in Canada. However, until recently, palivizumab, a monoclonal antibody, was the sole product available for RSV prevention in children and only indicated for infants meeting stringent criteria encompassing early gestational age and the presence of specific chronic conditions.^11^ In order to protect all infants from severe RSV disease, an extended half-life humanized monoclonal antibody with neutralizing activity was developed. Nirsevimab acts as a viral inhibitor by binding to the fusion protein on the surface of the RSV virus.^12^ Two clinical trials have demonstrated a statistically significant reduction in the incidence of medically-attended (MA) RSV LRTI after nirsevimab administration compared to placebo in preterm infants born during or entering their first RSV season, term and late preterm infants, and in children up to 24 months of age who remain vulnerable to severe RSV disease through their second RSV season due to being preterm or having chronic lung disease of prematurity or congenital heart disease.^13–15^ In Canada, nirsevimab is currently indicated for the prevention of LRTI in neonates and infants during their first RSV season and in children up to 24 months at high-risk for severe RSV disease in their second RSV season.^16^ In the US, the Advisory Committee on Immunization Practices (ACIP) recently provided a unanimous recommendation for the administration of nirsevimab to all infants aged <8 months born during or entering their first RSV season and for those 8-19 months at increased risk for severe RSV entering their second RSV season.^17^ Given the severity and seasonality of RSV, it is important to understand the cost and clinical impact of different implementation scenarios within the pediatric population to ensure optimal and cost-effective protection. We implemented an externally validated economic model comparing nirsevimab to the standard of care in Canada, utilizing various willingness-to-pay (WTP) thresholds to determine the optimized price per dose for nirsevimab in Canada for the reduction of medically-attended RSV infections and hospitalizations from a societal perspective.

## Methods

### Overview

A static decision tree model followed a cohort of Canadian newborn infants segmented into three categories based on risks for severe RSV: 1) preterm infants (<37 weeks gestational age [WGA]) eligible to receive palivizumab (“palivizumab-eligible”); 2) healthy preterm infants (<37 weeks wGA); and 3) infants born at term (≥37 wGA). The definition of palivizumab-eligible infants was based on criteria reported in the Canadian literature.^18^ Based on Canadian RSV surveillance data, the RSV season was assumed to increase in activity from November to the end of the following March.^16^ In the base case scenario, all infants born outside and inside the RSV season were administered nirsevimab during their first RSV season in the treatment arm (those born prior to the season received nirsevimab at the season’s start), with a select proportion of preterm infants receiving follow-up treatment in the subsequent season. Therefore, the newborn cohort was followed for two RSV seasons. The comparator arm was the standard of care (i.e., palivizumab), which only applied to a small proportion of infants meeting Canada’s strict eligibility criteria.^18^ The model simulated the patient trajectory of RSV infection with health outcomes, including RSV-related hospitalization, intensive care admission (ICU), mechanical ventilation (MV), emergency and general practitioner admissions (ER, GP), and RSV-related mortality. Direct and indirect costs associated with MA RSV LRTI were integrated into the model, which assumed the societal perspective. All mortality-related costs and outcomes were discounted at a rate of 1.5%, as recommended by national guidelines.^19^

The model integrated the following elements: population data (segmented by palivizumab-eligible, preterm, and term infants), seasonality of RSV, coverage and adherence rates, clinical profiles of therapies, RSV-related health outcomes (e.g., hospitalization, ICU, MV), outcome-related utility values, and outcome specific costs. In order to capture the seasonality of RSV, the implementation scenarios of nirsevimab by subgroup (e.g., born in-season, born out-of-season, palivizumab-eligible, preterm, term), and an accurate assessment of price per dose, the economic model required granular (monthly) data inputs. These monthly inputs included population birth, probability of RSV infection, and RSV-related health outcomes. A comprehensive list of parameters utilized in the model is presented in Table 1.

**Table 1.**
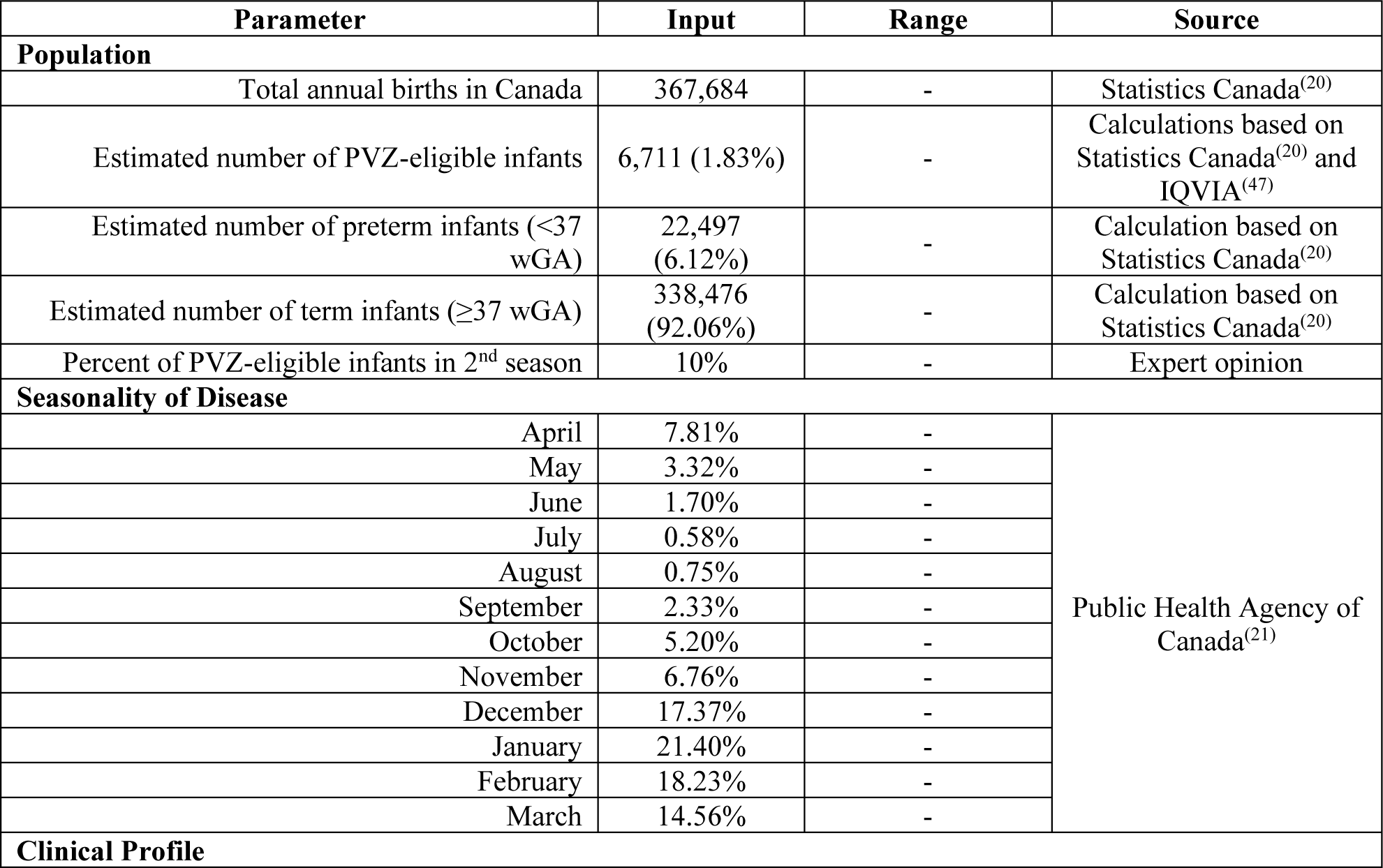
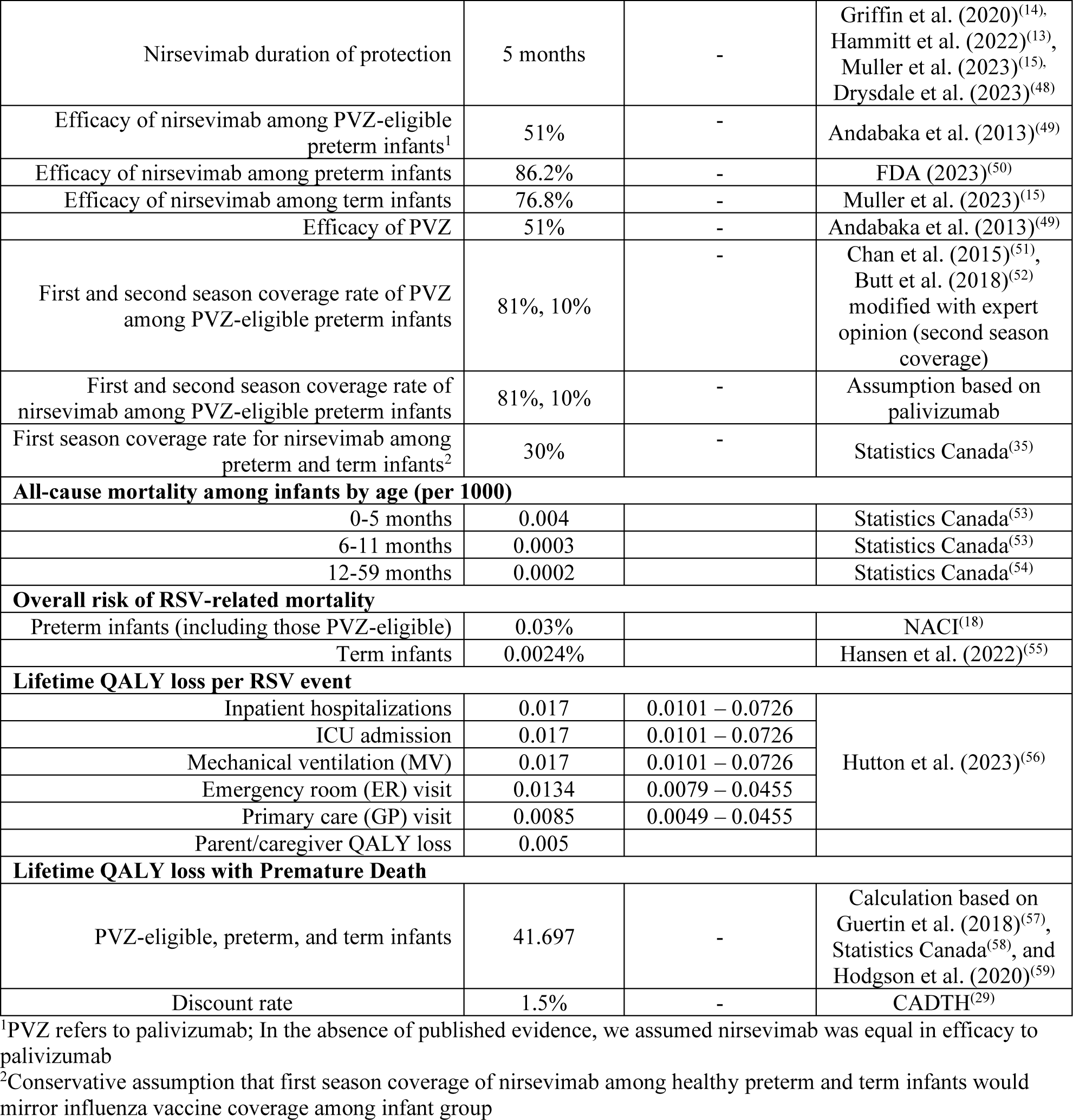
Key model inputs. Table 1 is a comprehensive list of all key model inputs, including population assumptions, risks associated with seasonality of RSV, clinical profiles of nirsevimab and palivizumab, mortality rates and lifetime QALY losses related to RSV.

### Health Outcome Parameters

The grey and peer-reviewed literature was systematically reviewed to identify ongoing and completed health economic, clinical, and epidemiological studies related to RSV and palivizumab. Population-based data, with the subgroups of interest and the monthly probability of infection, were derived from national statistics and surveillance data.^20,21^ All inputs for nirsevimab were derived from Phase 2b and Phase 3 studies, while the grey literature provided insights into palivizumab’s cost and clinical profile.^13–15,18,22^ Where possible, only up-to-date Canadian inputs were utilized to populate the model; however, in the complete absence of such data, US-specific inputs were adopted (e.g. data related to RSV-specific QALY loss and mortality outcomes). Data associated with RSV-related inpatient hospitalization, ICU admissions, and patients requiring MV were generated per infant subgroup, using a combination of average Canadian inputs and distributions of monthly US data (0-36 months age of infection). Informed by expert opinion, we assumed US-based monthly distributions as an acceptable proxy for generating “Canadianized” values per subgroup. Figure 1 illustrates the distributional conversion formula for generating Canadian values, utilizing an average of Canadian-specific inputs (see Table 2). Furthermore, for ER and primary care admissions, US-based monthly percentages (0-36 months age of infection) were utilized in the absence of monthly data for Canada.^23^

**Figure 1.**
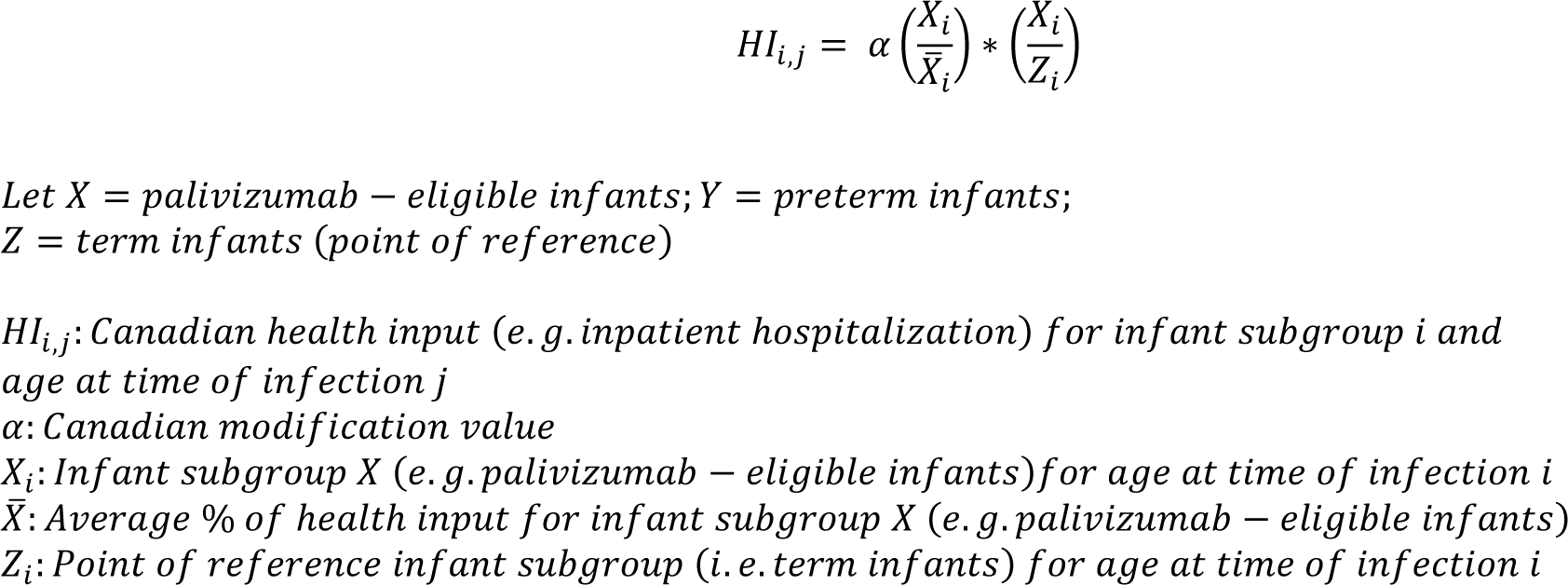
Modification formula for health-related inputs. The formula in Figure 1 was constructed to utilize average Canadian values and generate data points specific to health outcome, age of infection, and sub-group of interest, assuming a similar distribution of cases to the United States.

**Table 2.** Health-related inputs. Health-related inputs for RSV are presented in Table 2 by wGA and infant category risk for this disease.

| Inpatient hospitalization |  |  |  |  |
| --- | --- | --- | --- | --- |
| Age at time of infection (months) | PVZ-eligible infants | Preterm infants | Term Infants | Source |
| 0 | 13.18% | 3.32% | 1.89% | Feldes et al. (2015)<sup>(60)</sup><br>Hall et al. (2013)<sup>(61)</sup><br>IMPACT RSV Study Group (2018)<sup>(62)</sup><br>McLaurin et al. (2016)<sup>(28)</sup><br>Schanzer et al. (2018)<sup>(5)</sup> |
| 1 | 25.28% | 6.37% | 3.63% |  |
| 2 | 13.96% | 3.52% | 2.00% |  |
| 3 | 10.05% | 2.53% | 1.44% |  |
| 4 | 8.69% | 2.19% | 1.25% |  |
| 5 | 4.68% | 1.18% | 0.67% |  |
| 6 | 4.00% | 1.01% | 0.57% |  |
| 7 | 5.47% | 1.38% | 0.78% |  |
| 8 | 3.32% | 0.84% | 0.48% |  |
| 9 | 3.71% | 0.93% | 0.53% |  |
| 10 | 3.61% | 0.91% | 0.52% |  |
| 11 | 2.83% | 0.71% | 0.41% |  |
| 12-24 | 2.50% | 0.59% | 0.20% |  |
| 25-36 | 2.50% | 0.59% | 0.20% |  |
| Intensive Care Unit (ICU) |  |  |  |  |
| 0 | 43.36% | 60.35% | 18.81% | Arriola et al. (2020)<sup>(63)</sup><br>Mitchell et al. (2017)<sup>(64)</sup> |
| 1 | 43.36% | 60.35% | 18.81% |  |
| 2 | 43.36% | 60.35% | 18.81% |  |
| 3 | 19.66% | 15.43% | 13.95% |  |
| 4 | 19.66% | 15.43% | 13.95% |  |
| 5 | 19.66% | 15.43% | 13.95% |  |
| 6 | 11.42% | 9.28% | 10.31% |  |
| 7 | 11.42% | 9.28% | 10.31% |  |
| 8 | 11.42% | 9.28% | 10.31% |  |
| 9 | 11.42% | 9.28% | 10.31% |  |
| 10 | 11.42% | 9.28% | 10.31% |  |
| 11 | 11.42% | 9.28% | 10.31% |  |
| 12-24 | 18.67% | 31.26% | 10.92% |  |
| 25-36 | 18.67% | 31.26% | 10.92% |  |
| Mechanical Ventilation (MV) |  |  |  |  |
| 0 | 14.62% | 22.20% | 8.20% | Arriola et al. (2020)<sup>(63)</sup><br>Buchan et al. (2019)<sup>(27)</sup> |
| 1 | 14.62% | 22.20% | 8.20% |  |
| 2 | 14.62% | 22.20% | 8.20% |  |
| 3 | 5.69% | 6.80% | 1.82% |  |
| 4 | 5.69% | 6.80% | 1.82% |  |
| 5 | 5.69% | 6.80% | 1.82% |  |
| 6 | 5.69% | 4.72% | 1.82% |  |
| 7 | 5.69% | 4.72% | 1.82% |  |
| 8 | 5.69% | 4.72% | 1.82% |  |
| 9 | 5.69% | 4.72% | 1.82% |  |
| 10 | 5.69% | 4.72% | 1.82% |  |
| 11 | 5.69% | 4.72% | 1.82% |  |
| 12-24 | 1.37% | 1.13% | 2.73% |  |
| 25-30 | 1.37% | 1.13% | 2.73% |  |
| Emergency Room (ER) |  |  |  |  |
| 0 | 1.96% | 1.96% | 1.96% | Lively et al.<br>(2019)<sup>(23)</sup> |
| 1 | 6.42% | 6.42% | 6.42% |  |
| 2 | 7.24% | 7.24% | 7.24% |  |
| 3 | 10.52% | 10.52% | 10.52% |  |
| 4 | 11.60% | 11.60% | 11.60% |  |
| 5 | 7.13% | 7.13% | 7.13% |  |
| 6 | 8.18% | 8.18% | 8.18% |  |
| 7 | 5.61% | 5.61% | 5.61% |  |
| 8 | 5.56% | 5.56% | 5.56% |  |
| 9 | 5.56% | 5.56% | 5.56% |  |
| 10 | 4.04% | 4.04% | 4.04% |  |
| 11 | 5.56% | 5.56% | 5.56% |  |
| 12-24 | 5.30% | 5.30% | 5.30% |  |
| 25-36 | 5.30% | 5.30% | 5.30% |  |
| Primary Care (GP) |  |  |  |  |
| 0 | 8.52% | 8.52% | 8.52% | Lively et al.<br>(2019)<sup>(23)</sup> |
| 1 | 18.79% | 18.79% | 18.79% |  |
| 2 | 23.42% | 23.42% | 23.42% |  |
| 3 | 23.26% | 23.26% | 23.26% |  |
| 4 | 26.50% | 26.50% | 26.50% |  |
| 5 | 28.92% | 28.92% | 28.92% |  |
| 6 | 26.47% | 26.47% | 26.47% |  |
| 7 | 20.72% | 20.72% | 20.72% |  |
| 8 | 27.78% | 27.78% | 27.78% |  |
| 9 | 22.72% | 22.72% | 22.72% |  |
| 10 | 24.17% | 24.17% | 24.17% |  |
| 11 | 25.81% | 25.81% | 25.81% |  |
| 12-24 | 18.03% | 18.03% | 18.03% |  |
| 25-36 | 18.03% | 18.03% | 18.03% |  |

Abbreviation PVZ: palivizumab

RSV-related input parameters are provided in Tables 1 and 2.

### Economic Parameters

All costs are outlined in Table 3 with inflation adjustments using the Canadian Consumer Price Index (CPI), and adjusted to 2023 Canadian dollars. Although the model adopts a national scope, most RSV-specific costs within the past five years were from Ontario, Alberta, and Quebec-specific peer-reviewed sources due to availability.^24–27^ Furthermore, grey literature on the cost-effectiveness and recommended use of palivizumab provided overall support for model parametrization.^18,22^ Direct healthcare costs per RSV-related health outcome required the establishment of a costing matrix with an average cost per health outcome by subgroup (i.e. palivizumab-eligible, preterm, and term infants). ER, GP, and direct non-medical costs per hospitalization (i.e., travel, meals, childcare, and out-of-pocket expenses attributed to RSV-related hospitalization) were assumed to be uniform across subgroups and remained constant regardless of scenario analysis. In contrast, inpatient hospitalization, ICU, and MV costs required calculations based on different sources. The primary costing scenario (S_0_) was used for base case modelling; intended as a representation of a broader Canadian costing perspective, it was generated by averaging all scenarios (S1a, S1b, S2, S3, and S4, as described below). For the majority of the scenarios, the distribution of costs per subgroup were derived from McLaurin et al.^28^ For Scenario S1a and S1b, the recent Canadian literature from Rafferty et al. provided average RSV-attributable lab-confirmed hospitalization costs within 30 days and 365 days post-diagnosis by age group.^27^ Utilizing matrix algebra, the case-weighted cost for infants <1 year was applied to the distribution from McLaurin et al. to establish values within the costing matrix S1a and S1b.^28^ In Scenario 2 (S2), cost outcomes were based on Thampi et al. (2021), with cost differentials between palivizumab-eligible, preterm, and term infants but not across the different health outcomes (e.g., inpatient hospitalization, ICU, MV).^26^ Scenario 3 (S3), derived inputs from Buchan et al. (2019), and applied a similar costing approach as S2.^27^ Scenario 4 (S4) utilized the detailed costs from Papenberg et al. (2020), where direct and indirect RSV-related costs were sourced from multiple Quebec-specific resources (e.g. the Quebec government’s health insurance board Régie de l’assurance maladie du Québec [RAMQ], McGill University Health Centre, etc.).^25^

**Table 3.**
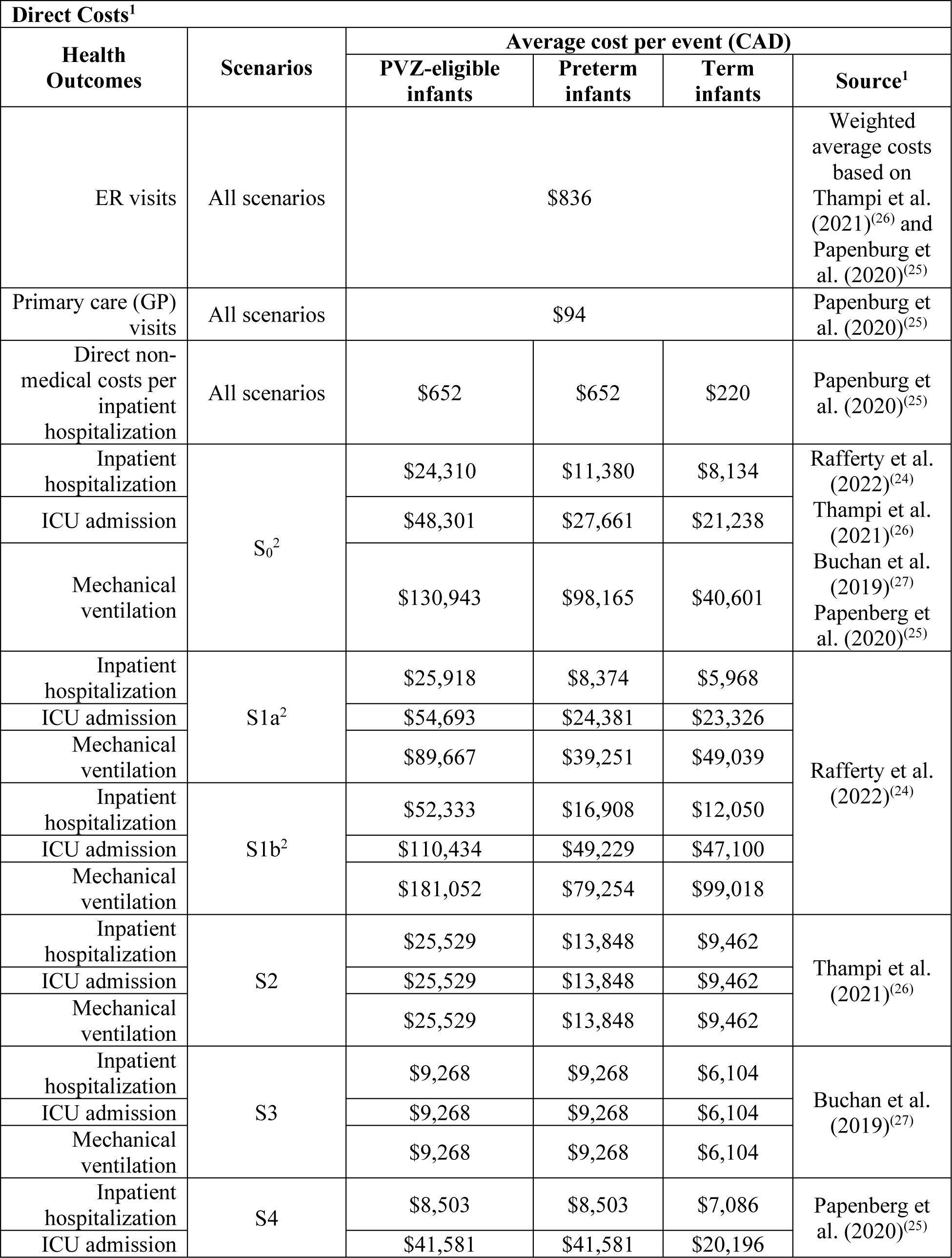
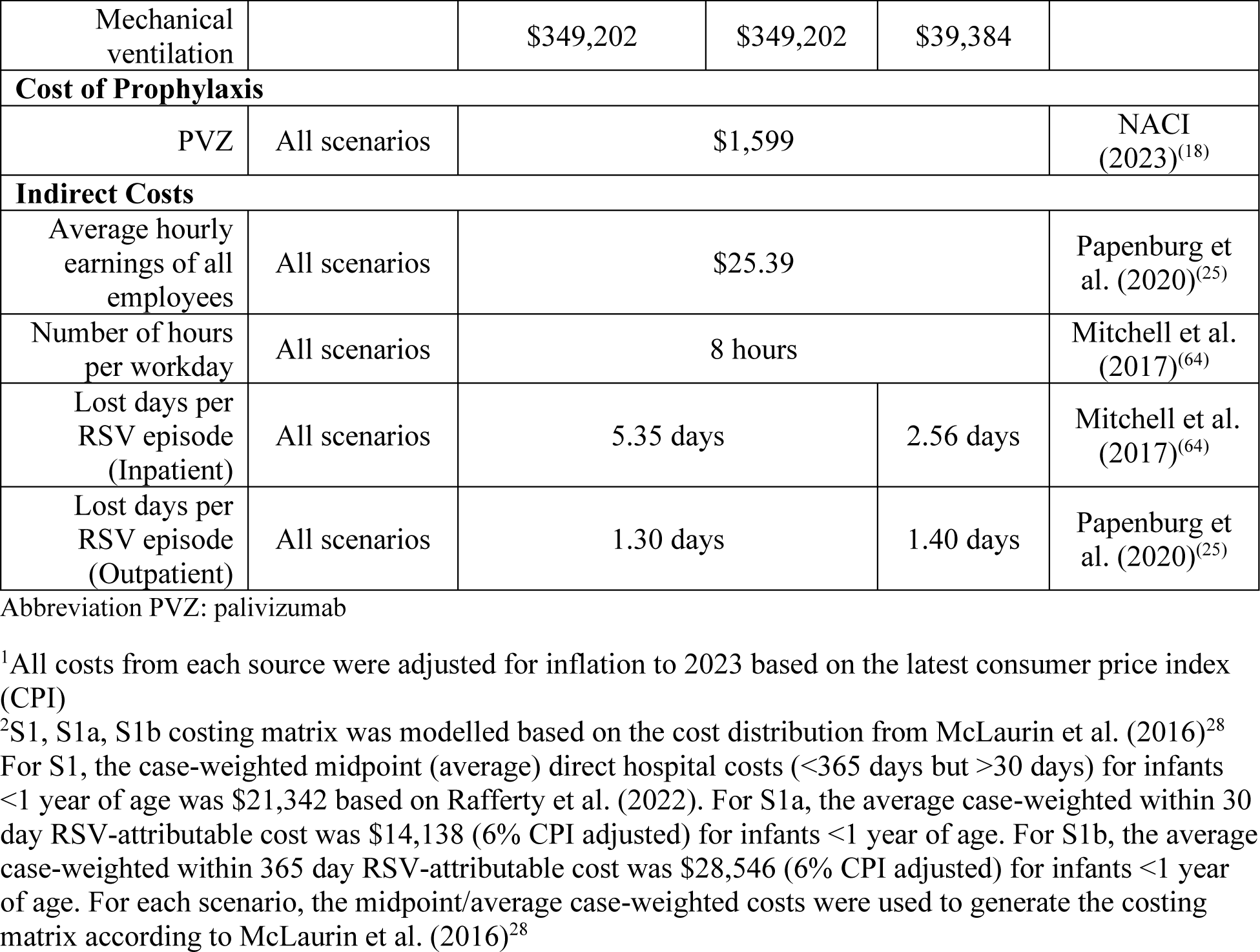
Cost-related inputs. Table 3 displays the RSV-related healthcare resource costs utilized in the model, stratified by infant risk for RSV.

| Direct Costs<sup>1</sup> |  |  |  |  |  |
| --- | --- | --- | --- | --- | --- |
| Health Outcomes | Scenarios | Average cost per event (CAD) |  |  |  |
|  |  | PVZ-eligible infants | Preterm infants | Term infants | Source<sup>1</sup> |
| ER visits | All scenarios | \$836 | | | Weighted average costs based on Thampi et al. (2021)<sup>(26)</sup> and Papenburg et al. (2020)<sup>(25)</sup> |
| Primary care (GP) visits | All scenarios | \$94 | | | Papenburg et al. (2020)<sup>(25)</sup> |
| Direct non-medical costs per inpatient hospitalization | All scenarios | \$652 | \$652 | \$220 | Papenburg et al. (2020)<sup>(25)</sup> |
| Inpatient hospitalization | S<sub>0</sub><sup>2</sup> | \$24,310 | \$11,380 | \$8,134 | Rafferty et al. (2022)<sup>(24)</sup> |
| ICU admission | | \$48,301 | \$27,661 | \$21,238 | Thampi et al. (2021)<sup>(26)</sup> |
| Mechanical ventilation | | \$130,943 | \$98,165 | \$40,601 | Buchan et al. (2019)<sup>(27)</sup><br>Papenburg et al. (2020)<sup>(25)</sup> |
| Inpatient hospitalization | S1a<sup>2</sup> | \$25,918 | \$8,374 | \$5,968 | Rafferty et al. (2022)<sup>(24)</sup> |
| ICU admission | | \$54,693 | \$24,381 | \$23,326 | |
| Mechanical ventilation | | \$89,667 | \$39,251 | \$49,039 | |
| Inpatient hospitalization | S1b<sup>2</sup> | \$52,333 | \$16,908 | \$12,050 | |
| ICU admission | | \$110,434 | \$49,229 | \$47,100 | |
| Mechanical ventilation | | \$181,052 | \$79,254 | \$99,018 | |
| Inpatient hospitalization | S2 | \$25,529 | \$13,848 | \$9,462 | Thampi et al. (2021)<sup>(26)</sup> |
| ICU admission | | \$25,529 | \$13,848 | \$9,462 | |
| Mechanical ventilation | | \$25,529 | \$13,848 | \$9,462 | |
| Inpatient hospitalization | S3 | \$9,268 | \$9,268 | \$6,104 | Buchan et al. (2019)<sup>(27)</sup> |
| ICU admission | | \$9,268 | \$9,268 | \$6,104 | |
| Mechanical ventilation | | \$9,268 | \$9,268 | \$6,104 | |
| Inpatient hospitalization | S4 | \$8,503 | \$8,503 | \$7,086 | Papenburg et al. (2020)<sup>(25)</sup> |
| ICU admission | | \$41,581 | \$41,581 | \$20,196 | |
| Mechanical ventilation | | \$349,202 | \$349,202 | \$39,384 | |
| Cost of Prophylaxis |  |  |  |  |  |
| PVZ | All scenarios | \$1,599 | | | NACI (2023)<sup>(18)</sup> |
| Indirect Costs |  |  |  |  |  |
| Average hourly earnings of all employees | All scenarios | \$25.39 | | | Papenburg et al. (2020)<sup>(25)</sup> |
| Number of hours per workday | All scenarios | 8 hours |  |  | Mitchell et al. (2017)<sup>(64)</sup> |
| Lost days per RSV episode (Inpatient) | All scenarios | 5.35 days |  | 2.56 days | Mitchell et al. (2017)<sup>(64)</sup> |
| Lost days per RSV episode (Outpatient) | All scenarios | 1.30 days |  | 1.40 days | Papenburg et al. (2020)<sup>(25)</sup> |

Abbreviation PVZ: palivizumab
<sup>1</sup>All costs from each source were adjusted for inflation to 2023 based on the latest consumer price index (CPI)
<sup>2</sup>S1, S1a, S1b costing matrix was modelled based on the cost distribution from McLaurin et al. (2016)<sup>28</sup> For S1, the case-weighted midpoint (average) direct hospital costs (<365 days but >30 days) for infants <1 year of age was \$21,342 based on Rafferty et al. (2022). For S1a, the average case-weighted within 30 day RSV-attributable cost was \$14,138 (6% CPI adjusted) for infants <1 year of age. For S1b, the average case-weighted within 365 day RSV-attributable cost was \$28,546 (6% CPI adjusted) for infants <1 year of age. For each scenario, the midpoint/average case-weighted costs were used to generate the costing matrix according to McLaurin et al. (2016)<sup>28</sup>

### Costing Scenarios

The price per dose optimization scenarios were based on four WTP thresholds (incremental cost per quality-adjusted life years [QALY]) of $40,000, $50,000, $70,000, and $100,000 per QALY, which represent the commonly accepted thresholds by North American health technology assessment (HTA) bodies, such as Canada’s Drug and Technology Agency (CADTH), Institut national d’excellence en santé en services sociaux (INESSS), National Advisory Committee of Immunization (NACI), and Institute for Clinical and Economic Review (ICER).^19,29–31^ The primary costing scenario (S_0_) attempted to represent a pan-Canadian perspective by averaging the costing matrices of all scenarios. Scenarios S1a and S1b utilized the average RSV-attributable lab-confirmed hospitalization costs within 30 days and 365 days (for those <1 year of age) to populate the costing matrix provided by McLaurin et al. Two additional scenarios (S2, S3) derived inputs from Ontario-specific sources with no assumptions regarding underlying cost distributions.^26,27^ In S2, costs were derived from Ontario’s RSV-focused population-based match cohort study.^26^ Costs for inpatient hospitalization, ICU, and MV among the preterm infants eligible for palivizumab were calculated based on the weighted mean adjusted costs for congenital heart disease (CHD), hemodynamically significant CHD, congenital lung disease (CLD), and birth between 22 and 28 wGA. Similar calculations were applied to preterm infants (29-35 wGA) and term infants (36-43 wGA or full-term with no comorbidities). Relative to the S_0_, S1a, S1b, and S2, the Ontario-based costing data in S3 was less granular, providing two average costs for young children ≤59 months with and without ≥1 comorbidity.^27^ S4 was based on Quebec-specific data providing the granular hospitalization costs attributed to RSV (e.g. pediatric unit, perinatal ICU, neonatal ICU, short-stay unit, ER) across a wide scope of cost outcomes ranging from hospital stay, administration, procedural, specialist, hospital discharge, outpatient resource use, productivity, and out-of-pocket auxiliary costs.^25^

### Implementation Scenarios

For all costing scenarios (S_0_, S1a, S1b, S2, S3, S4), analyses were conducted to explore the price per dose contingent on the scope of program implementation. The scope included four implementation scenarios: 1) all infants (i.e. palivizumab-eligible, preterm, and term) born within and outside of the RSV season (base case scenario); 2) palivizumab-eligible and preterm infants born within and outside of the RSV season (with the exclusion of term infants); 3) palivizumab-eligible and preterm infants born within and outside of the RSV season, and term infants 0-3 months entering their first RSV season and term infants born during the RSV season; and 4) palivizumab-eligible and preterm infants born within and outside of the RSV season and term infants born during the RSV season.

### Deterministic Sensitivity Analyses

A univariate sensitivity analysis was conducted to understand which variables significantly impacted the price per dose. The lower and upper bounds from the 95% confidence intervals (CI) were used where available; alternatively, we utilized a variance of ±20%.

## Results

### Price Per Dose

The price per dose of nirsevimab at the four WTP thresholds is presented in Table 4 for all costing and implementation scenarios, including the base case, which was consistently the least costly of the scenarios modelled. Using a WTP threshold of $40,000 per QALY, immunizing all infants born in and out of season would be cost-effective from a societal perspective at a PPD of $692 for the primary costing scenario (S_0_) and ranged from $634 to $769 depending on the source of costing input; at the highest WTP threshold of 100,000/QALY, the PPD increased to $894 (range: $836 to $971). Of all costing scenarios, S3, which derived its costing input from an Ontario-based RSV study that calculated costs for all children <59 months (stratified by presence or absence of at least one comorbidity) rather than more granular age groups, consistently yielded the lowest PPDs; the highest PPDs were generated by S1b, a scenario that used the average RSV-attributable lab-confirmed hospitalization costs within one year.

**Table 4.** Scenario-based price per dose ($CAD) of nirsevimab at various willingness-to-pay (WTP) thresholds. Price per dose (PPD) at the four studied WTP thresholds are presented in Table 4, per costing scenario and implementation scenario.

| Costing Scenarios<sup>1</sup> | Willingness-to-pay threshold (\$ CAD/QALY) | Implementation Scenarios | | | |
| --- | --- | --- | --- | --- | --- |
|  |  | All-infants born in & out-season (BASE CASE) | PVZ-eligible & preterm born in & out-season | PVZ-eligible & preterm born in & out-season, term infants (0-3 months) in 1st season and term infants born in-season | PVZ-eligible & preterm born in & out-season, term infants born in-season |
| **S<sub>0</sub>** | 40,000 | **\$ 692** | **\$ 3,767** | **\$ 874** | **\$ 1,117** |
| S1a | | \$ 677 | \$ 3,678 | \$ 854 | \$ 1,092 |
| S1b | | \$ 769 | \$ 3,875 | \$ 974 | \$ 1,235 |
| S2 | | \$ 660 | \$ 3,618 | \$ 829 | \$ 1,052 |
| S3 | | \$ 634 | \$ 3,565 | \$ 795 | \$ 1,018 |
| S4 | | \$ 721 | \$ 4,100 | \$ 918 | \$ 1,187 |
| **S<sub>0</sub>** | 50,000 | **\$ 726** | **\$ 3,827** | **\$ 908** | **\$ 1,149** |
| S1a | | \$ 711 | \$ 3,738 | \$ 888 | \$ 1,124 |
| S1b | | \$ 803 | \$ 3,935 | \$ 1,008 | \$ 1,267 |
| S2 | | \$ 694 | \$ 3,678 | \$ 863 | \$ 1,088 |
| S3 | | \$ 667 | \$ 3,625 | \$ 829 | \$ 1,049 |
| S4 | | \$ 755 | \$ 4,160 | \$ 952 | \$ 1,218 |
| **S<sub>0</sub>** | 70,000 | **\$ 793** | **\$ 3,946** | **\$ 976** | **\$ 1,213** |
| S1a | | \$ 778 | \$ 3,857 | \$ 956 | \$ 1,187 |
| S1b | | \$ 870 | \$ 4,054 | \$ 1,075 | \$ 1,330 |
| S2 | | \$ 762 | \$ 3,797 | \$ 930 | \$ 1,152 |
| S3 | | \$ 735 | \$ 3,745 | \$ 897 | \$ 1,113 |
| S4 | | \$ 823 | \$ 4,279 | \$ 1,020 | \$ 1,282 |
| **S<sub>0</sub>** | 100,000 | **\$ 894** | **\$ 4,126** | **\$ 1,077** | **\$ 1,308** |
| S1a | | \$ 879 | \$ 4,036 | \$ 1,057 | \$ 1,283 |
| S1b | | \$ 971 | \$ 4,233 | \$ 1,177 | \$ 1,426 |
| S2 | | \$ 863 | \$ 3,976 | \$ 1,032 | \$ 1,247 |
| S3 | | \$ 836 | \$ 3,924 | \$ 998 | \$ 1,209 |
| S4 | | \$ 924 | \$ 4,459 | \$ 1,121 | \$ 1,378 |

Abbreviation PVZ: palivizumab
<sup>1</sup>**S<sub>0</sub>**: costing scenario used in base case with average costs derived from S1a, S1b, S2, S3, and S4; S1a: costs derived from Rafferty et al. (2023) representing costs within 30 days post-RSV infection; S1b: costs derived from Rafferty et al. (2023) representing costs within 365 days post-RSV infection; S2: scenario based on costs derived from Thampi et al. (2020); S3: scenario based on costs derived from Buchan et al. (2019); S4: scenario based on costs derived from Papenberg et al. (2020)

Of all implementation scenarios, immunizing only palivizumab-eligible children and those preterm and born in and out of season generated the highest PPD for nirsevimab: $3,767 at a WTP threshold of $40,000/QALY and $4,126 at the highest threshold (i.e., $100,000/QALY).

### Clinical and Cost Impact

A comparison was conducted of the clinical and cost impact of administering nirsevimab to all infants (base case scenario) relative to the standard of care. Compared to current practice, the base case implementation strategy could avoid 18,249 RSV-related health outcomes (9.96%), including three inpatient deaths and 812 inpatient hospitalizations (Table 5). Use of the base case strategy also could avoid 4,071 ER visits, a reduction of 10.2%. The reduction of health outcomes is associated with a potential cost-savings of $26,324,578 (range of $19.7M to $34.9M for the additional scenarios utilizing short-term and long-term RSV-attributed hospitalization costs, respectively), of which 63.6% could be attributed to a reduction in direct costs, primarily derived from avoidance of hospitalization and ICU stays. The majority of both total outcomes avoided (99.7%), and resultant cost-savings (97.5%) are attributed to providing nirsevimab as RSV prophylaxis to otherwise healthy full-term and preterm children who are not eligible for palivizumab.

**Table 5.**
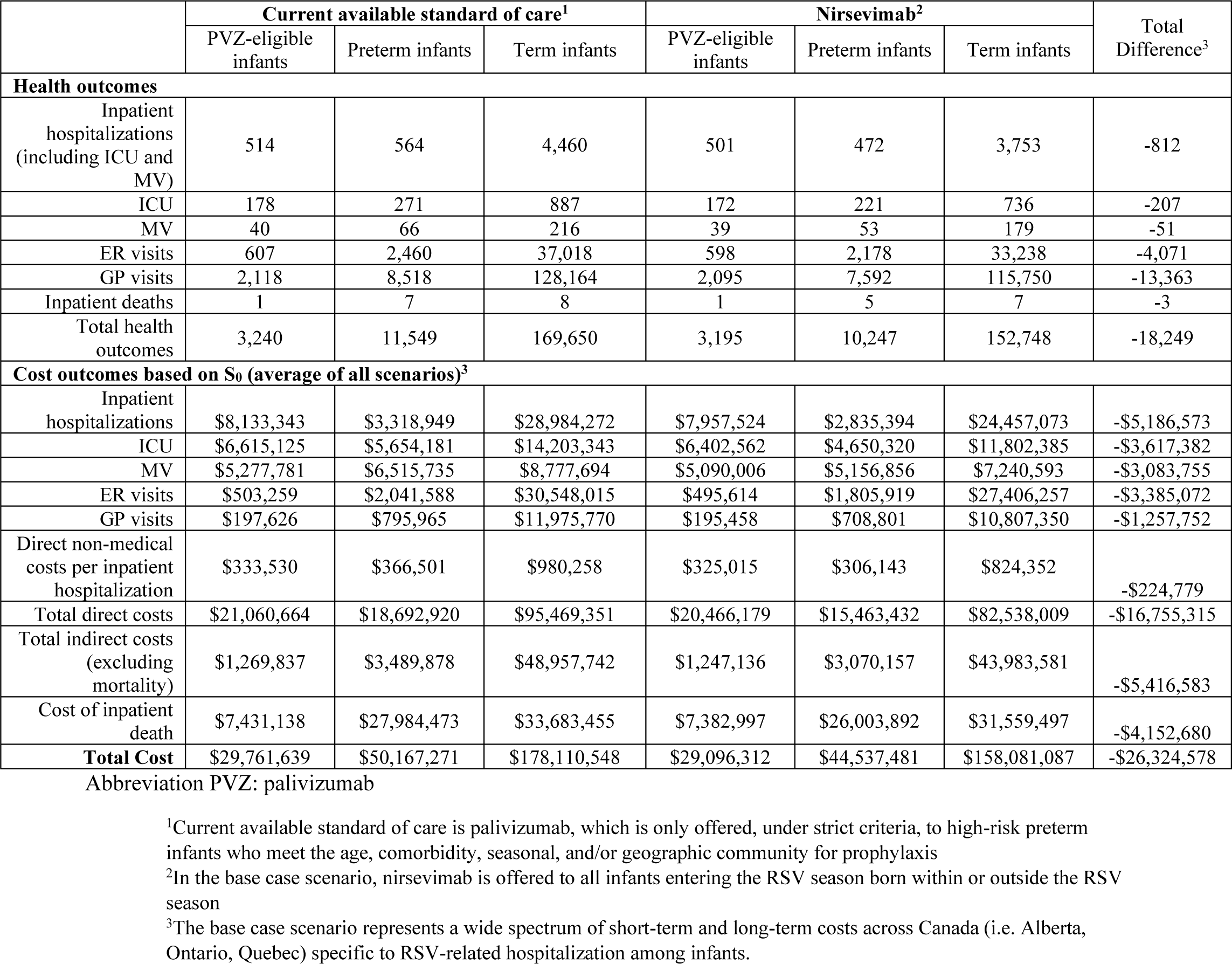
Comparative clinical and cost outcomes between existing therapy and nirsevimab for all newborn infants (born within and outside the RSV season) In Table 5, the base case scenario (nirsevimab is offered to all infants entering the RSV season born within or outside the RSV season) is compared to the standard of care (palivizumab is offered to a select segment of high-risk preterm infants). Clinical and cost outcomes are presented.

|  | Current available standard of care<sup>1</sup> |  |  | Nirsevimab<sup>2</sup> |  |  | Total Difference<sup>3</sup> |
| --- | --- | --- | --- | --- | --- | --- | --- |
|  | PVZ-eligible infants | Preterm infants | Term infants | PVZ-eligible infants | Preterm infants | Term infants |  |
| Health outcomes |  |  |  |  |  |  |  |
| Inpatient hospitalizations (including ICU and MV) | 514 | 564 | 4,460 | 501 | 472 | 3,753 | -812 |
| ICU | 178 | 271 | 887 | 172 | 221 | 736 | -207 |
| MV | 40 | 66 | 216 | 39 | 53 | 179 | -51 |
| ER visits | 607 | 2,460 | 37,018 | 598 | 2,178 | 33,238 | -4,071 |
| GP visits | 2,118 | 8,518 | 128,164 | 2,095 | 7,592 | 115,750 | -13,363 |
| Inpatient deaths | 1 | 7 | 8 | 1 | 5 | 7 | -3 |
| Total health outcomes | 3,240 | 11,549 | 169,650 | 3,195 | 10,247 | 152,748 | -18,249 |
| Cost outcomes based on S<sub>0</sub> (average of all scenarios)<sup>3</sup> |  |  |  |  |  |  |  |
| Inpatient hospitalizations | \$8,133,343 | \$3,318,949 | \$28,984,272 | \$7,957,524 | \$2,835,394 | \$24,457,073 | -\$5,186,573 |
| ICU | \$6,615,125 | \$5,654,181 | \$14,203,343 | \$6,402,562 | \$4,650,320 | \$11,802,385 | -\$3,617,382 |
| MV | \$5,277,781 | \$6,515,735 | \$8,777,694 | \$5,090,006 | \$5,156,856 | \$7,240,593 | -\$3,083,755 |
| ER visits | \$503,259 | \$2,041,588 | \$30,548,015 | \$495,614 | \$1,805,919 | \$27,406,257 | -\$3,385,072 |
| GP visits | \$197,626 | \$795,965 | \$11,975,770 | \$195,458 | \$708,801 | \$10,807,350 | -\$1,257,752 |
| Direct non-medical costs per inpatient hospitalization | \$333,530 | \$366,501 | \$980,258 | \$325,015 | \$306,143 | \$824,352 | -\$224,779 |
| Total direct costs | \$21,060,664 | \$18,692,920 | \$95,469,351 | \$20,466,179 | \$15,463,432 | \$82,538,009 | -\$16,755,315 |
| Total indirect costs (excluding mortality) | \$1,269,837 | \$3,489,878 | \$48,957,742 | \$1,247,136 | \$3,070,157 | \$43,983,581 | -\$5,416,583 |
| Cost of inpatient death | \$7,431,138 | \$27,984,473 | \$33,683,455 | \$7,382,997 | \$26,003,892 | \$31,559,497 | -\$4,152,680 |
| Total Cost | \$29,761,639 | \$50,167,271 | \$178,110,548 | \$29,096,312 | \$44,537,481 | \$158,081,087 | -\$26,324,578 |

Abbreviation PVZ: palivizumab
<sup>1</sup>Current available standard of care is palivizumab, which is only offered, under strict criteria, to high-risk preterm infants who meet the age, comorbidity, seasonal, and/or geographic community for prophylaxis
<sup>2</sup>In the base case scenario, nirsevimab is offered to all infants entering the RSV season born within or outside the RSV season
<sup>3</sup>The base case scenario represents a wide spectrum of short-term and long-term costs across Canada (i.e. Alberta, Ontario, Quebec) specific to RSV-related hospitalization among infants.

### Deterministic Sensitivity Analysis

The results of the univariate sensitivity analysis for the base case scenario (nirsevimab provided to all infants within and outside RSV season) at the $40,000 WTP threshold are displayed in the Tornado diagram in Figure 4. The factors resulting in the most variation in PPD were the discount rate, the variance of distribution of monthly RSV, the coverage rate of nirsevimab for infants born at term, and the cost of palivizumab. [Figure 4 here]

**Figure 2.**
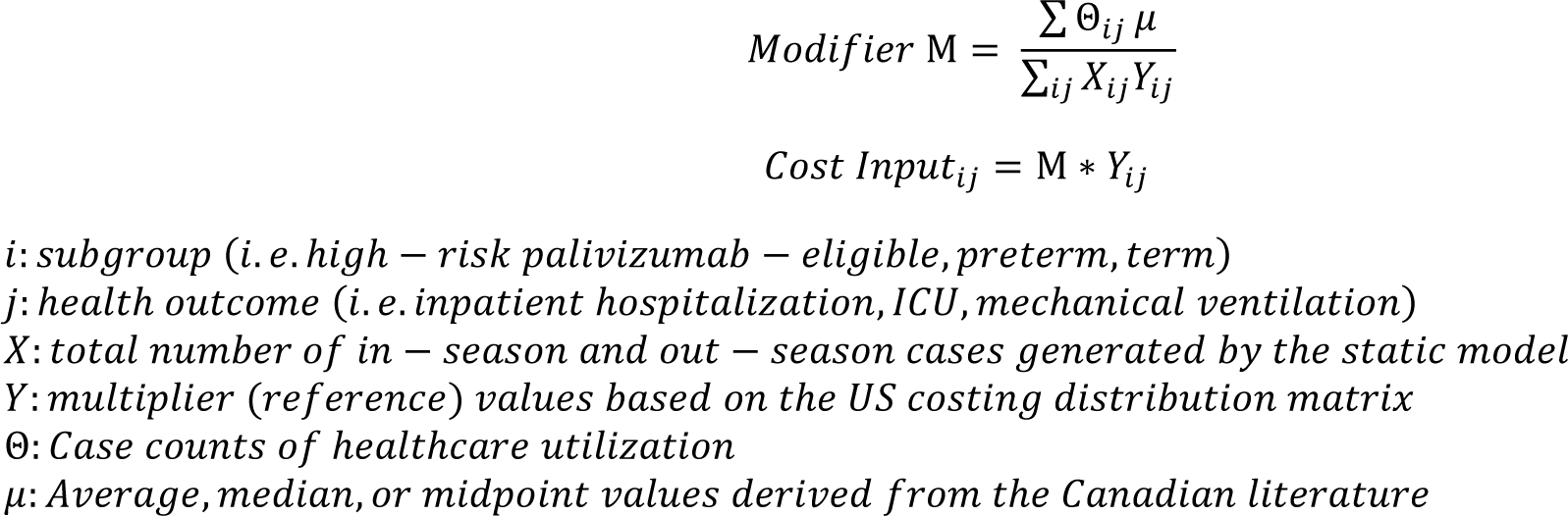
Modification formula for cost-related inputs [scenarios S1a, S1b based on Rafferty et al. (2023)] The formula in Figure 2 was constructed to utilize existing aggregate Canadian costs and generate specific outcome and sub-group specific costing data for RSV-related hospitalization, ICU admission, and mechanical ventilation, assuming a similar distribution of costs to the United States.

**Figure 3.**
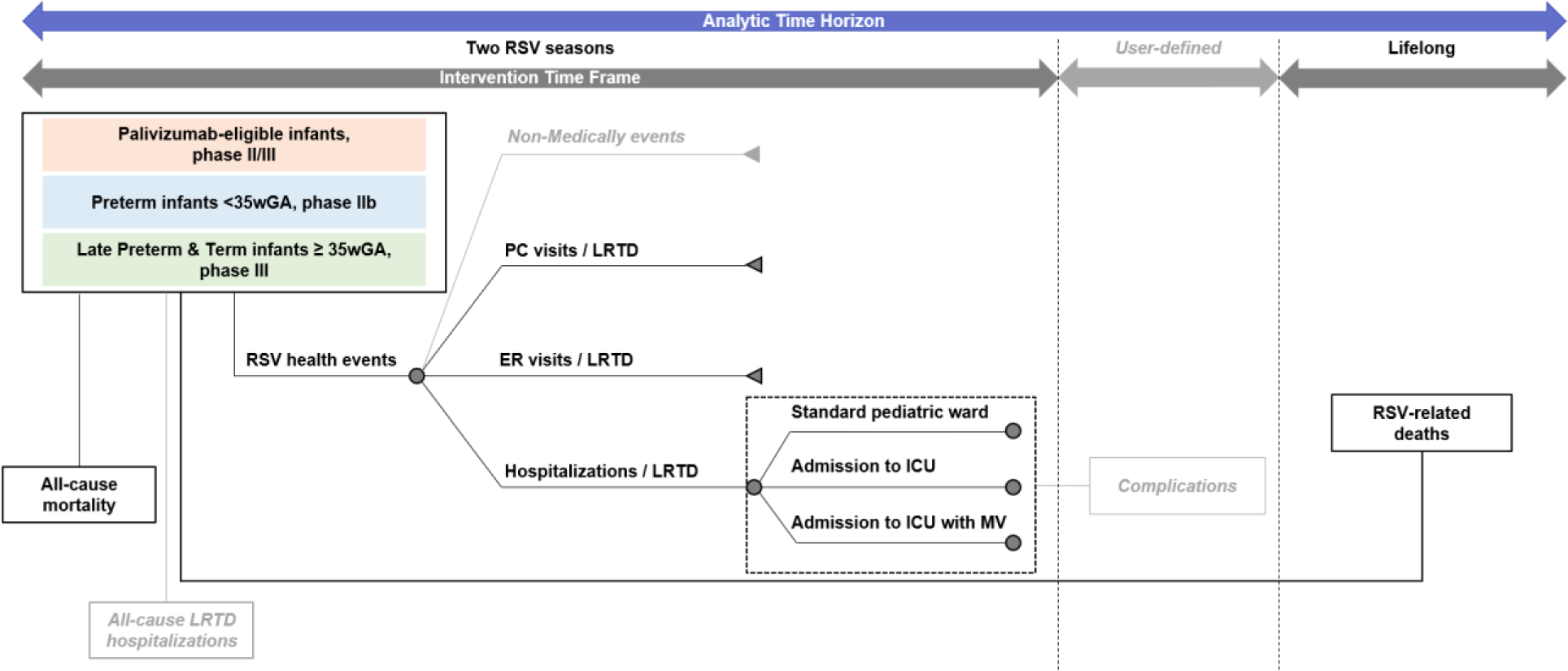
Model structure. Figure 3 provides a visual illustration of the static model with the corresponding subgroups, health outcomes, and timeframe of interest. The population includes infants less than two years of age, and it is divided into three subgroups to account for the differential individual risks of lower respiratory tract disease (LRTDs) and for the groups assessed in the nirsevimab clinical trials: Palivizumab-eligible infants, Preterm infants, and Late Preterm or Term infants. Infants experience different health events and require use of medical resources such as primary care (PC) and emergency room (ER) visits as well as hospitalizations. During the inpatient health state, infants might be in standard pediatric ward, admitted in intensive care unit (ICU) and requiring mechanical ventilation (MV). Non-medically events as referred to in this figure (in grey) are modelled in a scenario analysis, as well as all-cause LRTD hospitalizations, and complications following an RSV related hospitalization. Abbreviations: ER = emergency room; ICU = intensive care unit; LRTD = lower respiratory tract disease; MV = mechanical ventilation; PC = primary care; RSV = respiratory syncytial virus; wGA = week of gestational age.

**Figure 4.**
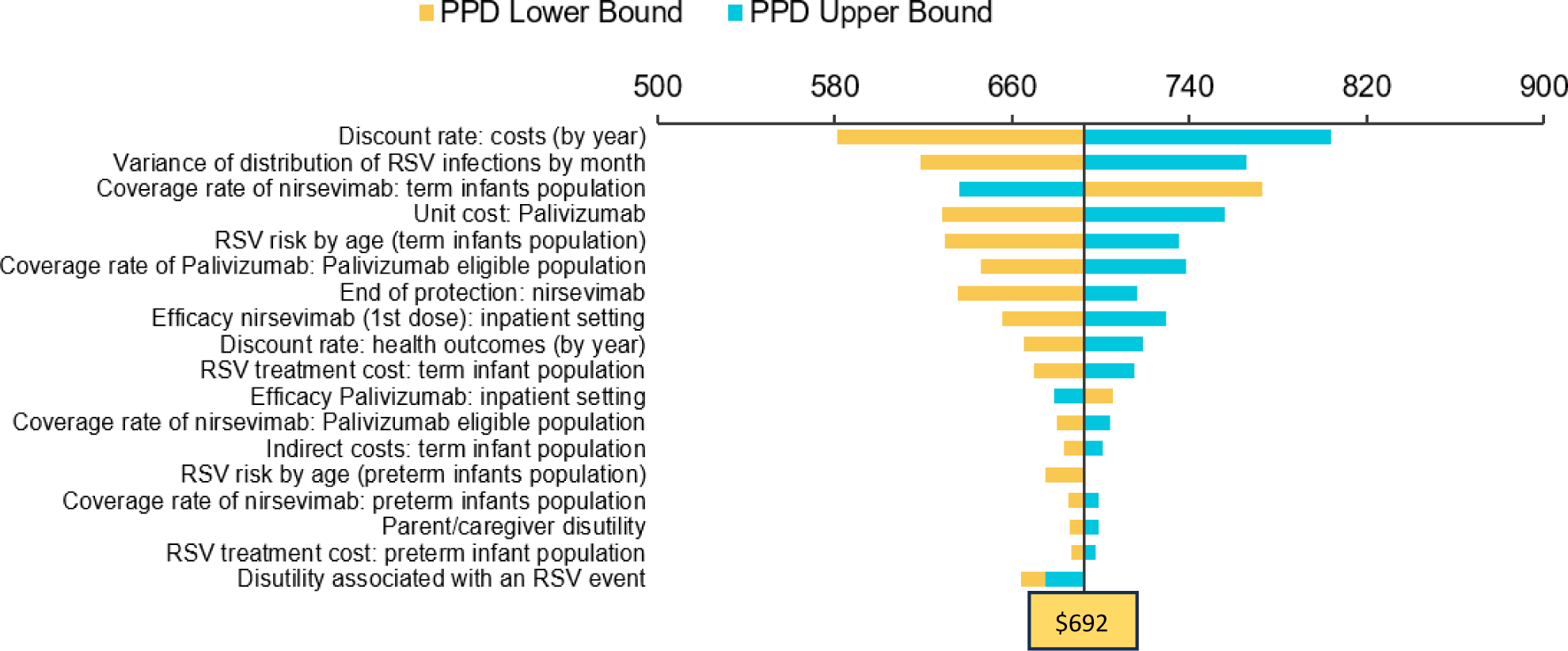
Deterministic sensitivity analysis for price per dose (PPD) The DSA was executed for all scenarios. Since each scenario only applies variations in the costing matrix, providing one DSA (base case) was considered appropriate to understand how each parameter affects the price per dose. Figure 3 **was provided to illustrate the base case scenario for all infants within and outside the RSV season at the $40,000 cost per QALY threshold.**

## Discussion

The approval of nirsevimab as RSV prophylaxis in Canada represents a valuable source of protection for infants and young children against a disease that is associated with significant morbidity and healthcare resource utilization. Our study demonstrates that compared to scenarios where only specific segments of the infant population receive nirsevimab, administering nirsevimab to all infants during their first RSV season was the most cost-effective scenario compared to standard care, yielding a PPD of $692 at a $40,000/QALY willingness-to-pay threshold. In comparison to current standard care, the base case could avoid 18,249 RSV-related health outcomes (reduction of 9.96%) for a cost savings of $26,324,578. Our model estimates that passive immunization of all infants with nirsevimab could prevent approximately 812 hospitalizations. The largest RSV-related hospitalization burden occurs in term and preterm children who are otherwise healthy and, therefore, not eligible for palivizumab. While premature infants are at the highest risk for severe RSV-related outcomes requiring hospitalization,^32,33^ they comprise only a small percentage of the birth cohort (and therefore a smaller overall health and cost burden) compared to healthy term and preterm infants.

Our sensitivity analysis revealed that increasing the discount rate, the variations in monthly disease incidence and the cost of palivizumab, and decreasing nirsevimab coverage rate (our model assumed a rate of 30%) would each result in a higher PPD for nirsevimab. Immunization programs are often sensitive to discounting since benefits may not be realized immediately, as with conventional curative therapies.^34^ Instead, there are often time delays between the time of dose administration and when disease is averted, and discount rates become more impactful. We based our coverage rate assumption on influenza vaccine uptake in the infant population;^35^ given the potential risk and severity of influenza as a respiratory infection, particularly in infants, we deemed this an appropriate proxy. As nirsevimab becomes routinely administered in the United States and elsewhere as part of RSV prophylaxis, a more informed coverage rate assumption will be possible.

Unlike many other diseases, the seasonality of RSV translates to a substantial variation in risk of infection from month to month. Previous studies have demonstrated that RSV-associated hospitalizations and healthcare costs were the highest for children born between September and February, who, therefore, had early exposure to the virus, with rates peaking in December and declining into April and after.^36–39^ Due to this monthly change in viral risk combined with granularity in palivizumab and nirsevimab’s current indications for use in Canada, we strived to utilize monthly data inputs from Canada in our model. Given the lack of such data and the common seasonality of RSV and similar incidence in infants in Canada and the United States,^40,41^ we opted to extrapolate US-based health outcomes and cost inputs and modify them using average Canadian values. Incorporating monthly data inputs was deemed critical to the ability of our model to estimate the clinical and cost impact of nirsevimab vs. standard care.

Our costing scenarios were informed by recent Canadian studies, which used different models and sources of data to evaluate the healthcare costs and burden of RSV in infants and young children^24–27^ Though the model by Rafferty estimates costs utilizing data from Alberta, it also incorporates national RSV positivity data to estimate the incidence and may be more likely to have widespread applicability across Canada than if a province-specific rate was used. Rafferty’s study was also unique in that it calculated outcomes at 30 days and 365 days post-infection, a valuable stratification given the progression of RSV illness and the changing need for healthcare resources. Thampi et al.’s model considered children under 24 months and examined direct medical costs per hospitalized child with RSV over a 12-month period. While Buchan et al.’s model did the same for children under five years of age in an earlier publication, Thampi et al. included attributable costs associated with ER visits, inpatient hospitalization, same-day surgeries, inpatient rehabilitation, physician and outpatient diagnostic and laboratory services. The model by Papenburg et al. utilized Quebec data for RSV and LRTI hospitalizations among infants younger than six months at the start of, or born during the RSV season, assessing costs up to 30 days discharge. Given that these studies vary considerably in model inputs and age group granularity, and utilize data from different provinces, our base case analysis utilized an average of these costing scenarios to capture a pan-Canadian perspective.

We calculated the optimal PPD of nirsevimab at various widely accepted WTP thresholds, as Canada does not adhere to one official cost-effectiveness threshold to inform national and provincial recommendations and decisions. At a $40,000/QALY threshold, our model generated a base case price per dose of $692, with a range of $634 to $769, depending on the source of costing input. Costing models from the United States Centers for Disease Control (CDC) and Prevention’s Advisory Committee on Immunization Practices (ACIP) have utilized a list price of $445 USD (approximately $600 CAD) per dose of nirsevimab.^42^ The Sanofi US health economic model for nirsevimab vs. standard care assumed a price of $500 USD (approximately $670 CAD),^42^ which is the approximate private market list price in the US.^43^ The current public CDC vaccine price list (per dose) indicates a $395 USD ($530 CAD) public price and a $495 USD ($665 CAD) private price regardless of the dose concentration. A recently published Canadian cost-effectiveness study established nirsevimab’s maximum price per dose (PPD) to range between $575 and $595 CAD based on a societal all-infant administration approach with consideration for RSV-related mortality, differing efficacy profiles (i.e. sigmoidal versus constant), 80%-100% product uptake, and a $50,000 per QALY willingness-to-pay (WTP) threshold.^44^ The analysis findings demonstrated that the larger the target infant population for nirsevimab administration, the greater the reduction in RSV burden; the maximum PPD may have been conservative given that there was no standard of care comparator for any subgroup of infants (i.e. palivizumab).^45^ Therefore, our model’s generated PPD for the base case scenario seems well-aligned with current prices of nirsevimab from the literature, with additional PPDs, which are higher in maximum price based on alternative implementation scenarios targeting select at-risk segments of the infant population.

Our analysis has several strengths, including the use of a range of hospitalization costing data and a costing matrix to incorporate hospitalization-related costs per level of RSV risk, the assessment of various immunization scenarios, the stratification of the infant population by gestational age at birth, and the inclusion of various medically-attended health outcomes and mortality. Furthermore, our model attempted to capture the variability of RSV-related burden and costs based upon the most up-to-date Canadian-specific economic literature. However, our model has several limitations. As stated earlier, we assumed that Canadian RSV health-related inputs would follow a US-based distribution; while this assumption was supported by the similar incidence rates of RSV in both countries, differences in healthcare delivery, practice patterns and healthcare-seeking behaviour may have led to variation that we did not consider. We also assumed that healthcare costs would differ between subgroups and adjusted our model’s hospitalization-related inputs according to the costing matrix; using US data to inform the matrix may have introduced differences in our model. However, Canadian literature suggests that hospitalization length of stay, need for ICU stay, and use of mechanical ventilation vary considerably based on wGA and underlying conditions, with the implication of significant differences in cost between risk subgroups by province.^25^ Based on our guidance from external experts, we deemed that integrating this into our model would enhance its accuracy. Additionally, our model assumed that infants with RSV infection would be immune for the rest of the study period and did not account for re-infection; the impact is likely low, given that previous studies report that re-infection is typically mild.^10^ Our model did not include the RSV-related costs associated with long-term sequelae such as asthma, wheezing, otitis media, and reduced pulmonary function incurred after hospitalization, which has been linked to RSV.^46^

Our study demonstrates that a passive immunization strategy for all infants with nirsevimab could be cost-effective at a WTP threshold of $40,000/QALY, provided the nirsevimab PPD is less than $692. This implementation scenario could result in significant reductions in the health and economic burden of RSV in Canada compared to the current standard of practice of administering palivizumab to certain at-risk infants only.

## Acknowledgements

Sanofi would like to acknowledge the input and feedback from various clinical and health economic experts from multiple advisory boards across Canada for the refinement and development of this costing model.

## Author Contributions

Study conception, design, analysis, interpretation and manuscript development were lead and supported by TS, JKHL, MG and AK; JW provided expertise in methodology and model validation. All authors reviewed and approved the final version of the manuscript and are accountable for all aspects of the work.

## Data Availability

Please contact the primary author for any inquiries pertaining to the model and input data.

